# New insights in Ulcerative Colitis Associated Gut Microbiota in South American Population: *Akkermansia* and *Collinsella*, two distinctive genera found in Argentine subjects

**DOI:** 10.1101/2020.07.29.20164764

**Authors:** Rosso Ayelén, Aguilera Pablo, Quesada Sofía, Cerezo Jimena, Spiazzi Renata, Conlon Carolina, Milano Claudia, Gregorio Iraola, Coluccio Leskow Federico, Penas-Steinhardt Alberto, S. Belforte Fiorella

## Abstract

**Background:** Globally, ulcerative colitis (UC) is the most common form of intestinal inflammation, which is believed to be the result of a deregulated immune system response to commensal microbiota in a genetically susceptible host. Multicellular organisms rely heavily on their commensal symbiotic microbiota, whose composition is closely related to intrinsic local characteristics and regulated or modified by environmental factors. In the present study we aim to describe the unknown gut microbiota of patients with UC in comparison with healthy individuals in order to find novel biomarkers for UC in our region.

**Methods:** We evaluated 46 individuals, 26 healthy non-UC controls and 20 UC patients, from the metropolitan area of Buenos Aires (BA), Argentina. Clinical features, biochemical tests and anthropometric measurements were determined. Fecal samples were collected and DNA was extracted for microbiota analysis. The hypervariable regions V3-V4 of the bacterial 16SR gene were sequenced using a MiSeq platform and sequences were analyzed using the QIIME2 environment. In addition, we looked for differential functional pathways using PICRUSt and compared the performance of three machine learning models to discriminate the studied individuals, using taxa and functional annotations.

**Results:** All UC patients were under clinical treatment with 70% of individuals in remission. We found no significant differences in gut microbiota richness or evenness between UC patients and non-UC controls (alpha diversity). Remarcably, microbial compositional structure within groups (beta diversity) showed differences: At the phylum level, *Verrucomicrobia* was overrepresented in controls while *Actinobacteria* was distinctive of UC patients; At the genus level *Bacteroides* and *Akkermancia* were significantly more abundant among controls while *Eubacterium* and *Collinsella* in UC patients. In addition, our results showed that carbohydrates metabolism was preponderant in UC patients, not observing a distinctive biochemical pathway for the healthy non-UC controls. Finally, in order to define a robust classifying method in our population, we evaluated the capability of three machine learning models to classify individuals. Our results reinforced the idea of functional compensation in microbiome communities, as models that used KEGG orthologs annotations had better capabilities than taxonomy to distinguish UC patients.

**Conclusions:** Our study provides new knowledge on the differences and similarities of the gut microbiota of UC patients as compared to non-UC controls of our population. This allows not only the association of local changes in gut microbial diversity with the pathology process, but also the future development of personalized nutritional and pharmacological therapies through the use of omic strategies describing the metagenomic profiles of the Argentine population.

## INTRODUCTION

Inflammatory bowel disease (IBD) represents a complex, polygenic chronic disorder of unknown etiology [1,2]. It is estimated that IBD is associated with industrialized countries, where the decrease in contact with microorganisms, parasites or their derivatives, promote an increase in the prevalence of chronic inflammatory diseases. This is consistent with the hygiene hypothesis that suggests that lack of early childhood exposure to pristine microbial conditions may increase the individual’s susceptibility to disease [3–5]. In particular IBD includes two main phenotypes: Crohn’s disease (CD) and Ulcerative Colitis (UC). However, between 10-17% of IBD patients do not have a definitive diagnosis of CD or UC, a phenomenon known as “inflammatory bowel disease unclassified” (IBDU) [6] UC manly comprises the rectum, affecting contiguously and symmetrically the colon being more severe distally. Dependent on the colonic segments involved, UC extent can be classified as proctitis, left-sided colitis, or extensive colitis [7]. Disease activity classification is based on the number of daily stools and the presence (or absence) of inflammation signs. Crohn’s colitis is not continuous or symmetrical, and usually does not involve the rectum [8,9]. CD is also associated with intestinal granulomas, strictures and fistulas, which are not common findings in UC [1]. In CD inflammation is often transmural, whereas in UC is typically confined to the mucosa. As eventually CD could compromise the colon and share UC pathognomonic manifestations, it’s diagnosis is often confused despite being different diseases. Colonic CD and UC need to be distinguished by differences in genetic predisposition markers, risk factors and clinical features. Globally, UC is the most common form of intestinal inflammation, which is believed to be the result of a deregulated immune system response to commensal microbiota in a genetically susceptible host [1,2,10]. This entity can be identified months or years after the first appearance of symptoms, requiring clinical, serological, radiological, endoscopic and histological information to define its prognosis and treatment.

Multicellular organisms, such as humans, rely heavily on their commensal symbiotic microbiota. This heterogeneous community is composed by microbial groups such as viruses, bacteria, archaea, fungi and other eukaryotes found in multiple body niches, such as intestine, skin, vagina, mouth, etc. [11]. In particular, the human gut microbiome outnumbers human cells and expresses more genes than those present in our genome [12,13]. These complex communities of microorganisms mediate physiologically important chemical transformations playing a key role in recovering energy and nutrients from diet as well as promoting ion absorption at the colon level [12,13]. It also provides protection against exogenous microorganisms, preventing the invasion of the intestinal mucosa by pathogenic microbes. The use of antibiotics or drugs, stress exposure, or dietary habits, among other factors, could break the ecological balance generating intestinal dysbiosis and promoting the overgrowth of pathobionts [14]. It is essential to assess the composition of the gut-associated microbiome in the context of UC, as certain groups of organisms may alter the communication between the immune system and commensal microbes, triggering an exacerbated response in the intestinal mucosa [15,16].

Little is known about the human microbiome of South American populations [17–19]. Reports of gut dysbiosis processes associated with UC in this region are almost null, despite reports of a rapid increase in the incidence of IBD in South America [3]. So far, most of the available literature on the study of the gut microbiome associated with UC points to developed countries, which differ both in the genetic background and in several environmental factors from our population [20]. In particular only three articles have been published studying the gut microbiome of different Argentine populations, two of them of our authorship. The first local pilot study reported the most relevant and abundant bacteria in samples from different body regions in individuals from Rosario, Santa Fe (Argentina) [21]. We recently published the microbial gut diversity of Buenos Aires (BA), being the first local report in this area [22]. Since BA is the second most populated agglomeration in South America and the southern hemisphere (with a large genetic and cultural component of European immigration interacting with local indigenous people), we compared BA microbiota analysis with different 16S rRNA gene sequence data sets. In particular, Santiago de Chile’s population turns out to be the closest to BA’s, principally due to the presence of *Verrucomicrobiales* of the genus *Akkermansia*. Additionally, we recently demonstrated gut dysbiosis in psoriasis patients from BA, suggesting a role in psoriasis pathophysiology. Furthermore, we developed a Psoriasis-Microbiota Index (PMI) with the potential to discriminate between psoriasis patients and controls across different populations, which could be used as a risk estimator biomarker in the clinical field [23].

In the present work we characterized the unknown gut microbiota associated with UC patients in the BA Metropolitan area and revealed differences in the intestinal microbiome between non-UC control and UC patients. Given the absence of studies of gut dysbiosis phenomena associated with complex pathologies with an inflammatory component in our region, the study of metagenomic biomarkers associated with UC in the Argentine population is of great interest. Additionally, we analyzed the general metabolic functions associated with microbial profiles. As a whole, our study provides new knowledge on the differences and similarities of the gut microbiota of UC patients compared to non-UC controls of the South American population. This characterization allows not only the association of local changes in gut microbial diversity with the pathology process [24,25], but also the future development of personalized nutritional and pharmacological therapies through the use of omic strategies describing the metagenomic profiles of the Argentine population.

## MATERIALS AND METHODS

### Ethics statement

This study received approval by the Ethics Committees of Hospital Español of BA (for non-UC control population) and Hospital Nacional Prof. Alejandro Posadas (for UC patients), according to local regulations and Helsinki declaration. Written informed consent was obtained from all study participants.

### Selection of participants and environmental data

This cross-sectional study recruited unrelated individuals, including consecutive UC patients attending the Gastroenterology Service of the Posadas Hospital. non-UC controls were selected according to age and body mass index (BMI) in order to match the patient population characteristics (Table1), considering the same geographical location for all participants. The exclusion criteria established for both non-UC controls and UC patients, considered individuals who have not received antibiotic therapy in the last 6 month, subjects on extreme diets (e.g. macrobiotics, vegans), surgical intervention in the gastrointestinal tract (gastrectomy, bariatric surgery), pregnancy, digestive neoplasias, patients on renal replacement therapy, transplant recipients and HIV infected. Additional exclusion criterias for non-UC controls were the presence of IBD or Irritable bowel syndrome (IBS), family history of IBD in first degree relatives, immunological disorders, hypertension, fatty liver disease, diabetes mellitus, malignancy background or any other serious internal disease, and alcohol abuse.

### General diagnosis and UC activity

Medical specialists apply ECCO guidelines for the diagnosis, treatment and surveillance of UC patients. Diagnosis was defined by combined studies of endoscopy, histology, serology and clinical data. Patients with UC were characterized based on Truelove & Witts for clinical activity [26] and Mayo Score for endoscopies, as follows: normal, mild disease (erythema, decreased vascular pattern, mild friability), moderate disease (marked erythema, lack of vascular pattern, friability, erosions), or severe disease (spontaneous bleeding, ulceration) [27,28]. Physicians make a complete study of these parameters guides in order to apply specific treatment either for activation states of UC nor to define patient’s remission, that is, stool frequency ≤ 3/day, no rectal bleeding, normal endoscopy and absence of histological acute inflammatory infiltrate confirming quiescent course of disease and mucosal healing [6]. To assess the general information about participants anthropometric measurements (height, weight, and waist circumference) and blood pressure were determined by standardized protocols. BMI was calculated as weight (kg)/[height(m)]^2^. After a 12-hour overnight fast, venous blood samples were obtained from volunteers for further analyses. Fasting plasma glucose (FPG), creatinine, total cholesterol, triglycerides, LDL-C, and HDL-C were determined by enzymatic methods in serum samples using standardized procedures. Hemoglobine, platelets, albumin and C-Reactive-Protein (CRP) were measured in order to determine UC severity. All biochemical measurements were performed at the Hospital Posadas Laboratory, BA, Argentina. Given the low predictive nature of serological data in UC patients, anti-Saccharomyces cerevisiae / antineutrophil cytoplasmic antibodies (ASCA/ANCA antibodies) were determined by immunofluorescence and immunoenzymatic standard techniques only in those patients who required it to define the diagnosis. Likewise, patients undergo annual video colonoscopies to monitor the pathology, as well as histopathological analysis of biopsy samples taken for this purpose.

### Sample collection and DNA extraction

Each participant was given a written protocol for sample collection, which considered the introduction of approximately 5g of stool into a sterile wide-mouth tube containing a teaspoon in the lid. DMSO/EDTA/saturated sodium chloride buffer was used to preserve the samples at both room and subzero temperatures, preventing freezing and cell damage as described [22,29]. The samples were aliquoted and stored at -80°C until use. DNA extraction was performed using QIAamp PowerFecal DNA Kit following the manufacturer’s instructions. The concentration and purity of the nucleic acids were determined by spectrophotometry in NanoDrop ND-1000 (NanoDrop Technologies, Wilmington, DE, USA).

### 16S bacterial rRNA fragment NGS

To amplify the 16S rRNA gene fragments of gut microbiota, 30 ng of purified DNA were used, and V3-V4 hypervariable regions of the bacterial 16S gene were amplified using 337F/805R primers. All segments of the variable regions of the 16S rRNA gene were normalized and multiplexed in a single tube. Sequencing was carried out using a MiSeq sequencer performing the synthesis sequencing methodology. Libraries were sequenced in the 5’ and 3’ directions (paired end mode), ensuring 300-500 bp long sequences and ∼ 150,000 average coverage for taxonomic identification.

### Sequence analysis and comparison of microbial communities

Sequences generated were analyzed using Quantitative Insights Into Microbial Ecology (QIIME2) version 2019.7 software package [30]. We used a sub-operational-taxonomic-unit approach “Deblur”, which uses error profiles to obtain putative error-free sequences from Illumina sequencing platforms [31]. To control variation in sequencing coverage, the data was rarified at a depth of 28.835 sequences per sample. Three α-diversity measures were tested with general linear model: (i) Shannon diversity, which defines the total number of species (species richness) weighted for their relative abundances (species evenness); (ii) Faith’s phylogenetic diversity, to assess the amount or proportion of branch length in a phylogenetic tree that leads to different organisms (species richness); and (iii) Simpson’s index, to represent the probability of two randomly picked elements of a sample being part of the same Operational Taxonomic Unit (OTU). In order to obtain a phylogenetic tree for diversity computation, we used Qiime2’s fragment-insertion [32] to phylogenetically place the sub-OTU sequences into the reference Greengenes 13.8 database with 99% identity tree [33]. To assess diversity variation in the bacterial community, we calculated β-diversity using robust compositional Aitchison distance [34]. We investigated differences in sub-OTU abundance between non-UC controls vs UC patients using the analysis of composition of microbiomes (ANCOM) framework [35] and ANOVA-Like Differential Gene Expression (ALDEx2) [36].

We follow a standard pipeline of Phylogenetic Investigation of Communities by Reconstruction of Unobserved States (PICRUSt), implemented in QIIME2, to generate Kyoto Encyclopedia of Genes and Genomes (KEGG) pathway profiles, as previously described [37]. To determine significant differences of KEGG Orthologs (KO) between groups we used ALDEx2. We performed three machine learning models using KEGG orthologs, Genus and the combination of both to classify non-UC controls and UC patients with k-fold cross validation Random Forest method implemented in the caret R package [38]..

### Data accession

Raw sequences of 16S rRNA gene reported in this article were deposited in NCBI Short Read Archive (SRA) and are accessible under the accession number PRJNA646271.

## RESULTS

### Characteristics of the studied population

All participants were 47 years old average, mostly overweight and from the same geographical location belonging to the metropolitan area of Buenos Aires (Table 1). All UC patients were under chronic medical treatment as they had previously been diagnosed. They all received mesalazine, being steroids or azathioprine in some cases used. None of the UC patients are undergoing biological treatment (Table 2).

**Table 1:** Population general description.

| Groups |  | UC | non-UC controls | p |
| --- | --- | --- | --- | --- |
| General descriptions | Overall subjects | n=20 | n=26 | - |
|  | Female % | 60 | 57,69 | ns |
|  | Male % | 40 | 42,31 | ns |
| | Mean age, years ± (SD) | 46,65 ± (17,81) | 48,69 ± (18,82) | ns |
| | Mean Height, m ± (SD) | 1.61 ± (0,085) | 1.64 ± (0,08) | ns |
| | Mean Weight, Kg ± (SD) | 69,10± (11,00) | 75.64 ± (15,01) | ns |
| | BMI ± (SD) | 26,60 ± (4,09) | 28,12 ± (5,29) | ns |

*No significant (ns). Variables were assessed by the Mann-Whitney U test.*

**Table 2:** UC patients characteristization.

| Groups |  | Total |
| --- | --- | --- |
| | | mean ± (SD) |
| Biochemical data | Haemoglobin, g/dL | 13,74 ± (1,41) |
| | us-CRP, mg/L | 0,35 ± (0,26) |
| | Albumin, g/dL | 4,55 ± (0,34) |
| | Platelets, $\times 10^3$/ml | 238,1 ± (58,02) |
| | FPG, mg/dL | 92,40 ± (13,36) |
| | Creatinine, mg/dL | 0,77 ± (0,16) |
| | Triglycerides, mg/dL | 92,4 ± (49,33) |
| | Total cholesterol, mg/dL | 177,2 ± (43,04) |
| | LDL-C, mg/dL | 109,1 ± (42,50) |
| | HDL-C, mg/dL | 56,87 ± (13,44) |
|  |  | n (%) |
| Autoimmunity | ASCA + | 0 (0) |
|  | ANCA + | 2 (10) |
| UC therapy | Mesalazine | 20 (100) |
|  | Steroids | 1 (5) |
|  | Azathioprine | 4 (20) |
|  | Rectal budesonide | 5 (25) |
|  | Infliximab/Adalimumab | 0 (0) |
| Lesion localization | E1 proctitis | 1 (5) |
|  | E2 left-sided colitis | 8 (40) |
|  | E3 extensive | 11 (55) |
| Clinical activity | Remission | 14 (70) |
|  | Mild | 6 (30) |
|  | Moderate | 0 (0) |
|  | Severe | 0 (0) |
| Endoscopic Mayo score | Normal | 10 (50) |
|  | Mild | 7 (35) |
|  | Moderate | 1 (5) |
|  | Severe | 2 (10) |
| Histology activity | Quiescent | 10 (50) |
|  | Inflammatory infiltrate | 10 (50) |

*Ultra sensitive C-Reactive Protein: us-CRP, Fasting plasma glucose: FPG, symbol + of ASCA and ANCA shows presence of autoantibodies, n means the number of patients accompanied by their percentages*

Blood samples were taken by the responsible professionals during recruitment in order to determine biochemical variables and autoantibodies. These complementary parameters are useful for monitoring UC patients, since they can usually have low hemoglobin due to intestinal bleeding, hyperplatelet, low albumin levels and high ultra sensitive C-reactive protein (us-CRP) indicating inflammation and tissue damage. As shown in Table 2, we found no abnormal values among the UC patients in these parameters.

In relation to the collected clinical data, most of the UC patients suffered from extensive or left-sided colitis, while it was observed one proctitis case. Notably our population had most of the colon compromised, observing colonoscopic alterations from the rectum to the descending colon. In order to define our patients states, clinical, endoscopic and histological activity of our population were evaluated. These three parameters were considered since the disease may not manifest itself clinically but may be reflected in the endoscopic and histological examinations. In this sense, 70% of UC patients were in remission and 30% presented clinical activity classified with May score as mild phenotypes. Interestingly, 50% of the sick patients showed the presence of inflammatory infiltrate in biopsy samples (Table 2).

### Microbial composition

Low quality reads and chimera sequences were filtered and denoised from the raw data with Deblur, eventually producing an average of 43.209 reads per sample. These reads corresponded to 1671 sub-operational taxonomic units (sub-OTUs) and identified 134 genera of Greengenes reference database. There was no significant difference between the number of sub-OTUs in UC patients and non-UC controls (p-value = 0.830). The most abundant phylums in non-UC controls and UC patients were *Bacteroidetes, Firmicutes, Proteobacteria, Actinobacteria* and *Verrucomicrobia*, last two showing significant differences (Fig. 1-A and Table 3).

**Table 3:** Distribution of phylas.

| Phylum | % non-UC controls (SD) | % UC (SD) | p-value |
| --- | --- | --- | --- |
| **Bacteroidetes** | **65,1 (0,8)** | **49,9 (1,17)** | **0,83** |
| **Firmicutes** | **28,2 (0,38)** | **42,7 (0,77)** | **0,051 ·** |
| **Proteobacteria** | **4,20 (0,11)** | **3,9 (0,39)** | **0,85** |
| **Actinobacteria** | **1,07 (0,08)** | **2,80 (0,11)** | **0,0003 \*** |
| **Verrucomicrobia** | **1,12 (0,10)** | **0,31 (0,04)** | **0,01 \*** |
| **Fusobacteria** | **0,11 (0,02)** | **0,02 (0,004)** | **0,65** |
| **Synergistetes** | **0,01 (0,002)** | **0,23 (0,05)** | **0,80** |
| **Lentisphaerae** | **0,13 (0,009)** | **0,02 (0,002)** | **0,15** |
| **Tenericutes** | **0,05 (0,005)** | **0,08 (0,009)** | **0,51** |

*Average and standard deviation of percentage of taxonomic composition in non-UC controls and UC patients. P-value is the Benjamini-Hochberg corrected P value of the Wilcoxon test obtained from ALDEx2. \* = p-value < 0.05, · = p-value < 0.1*

**Figure 1.**
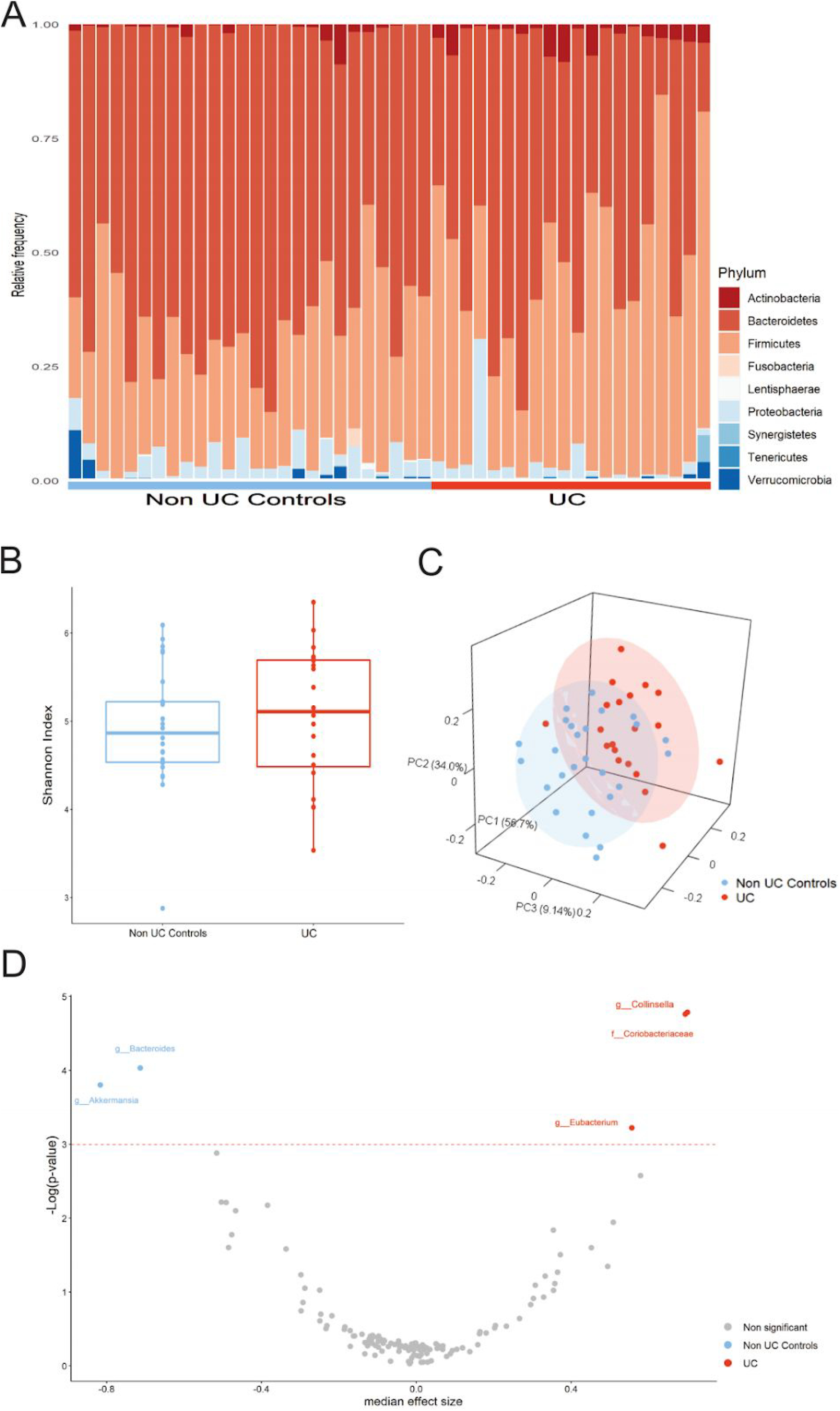
**A)** Microbiome taxonomic composition in non-UC controls and UC patients. Relative abundance at phylum level. Each column represents one fecal sample and different colors indicate different phylum in the microbiota composition. **B)** Alpha Diversity measure (Shannon diversity index) for non-UC control and UC patients. **C)** Beta diversity plot using robust compositional Aitchison distance between samples in our study with ellipses representing the 80% confidence interval of each group. **D)** Volcano plot of the differentially abundant genera between non-UC controls and UC patients. Red dash lines represent significance threshold (p-value = 0.05). Blue represents significant features more abundant in non-UC controls and red represents significant features more abundant in UC patients. In gray non significant features are represented.

### Alpha and Beta diversity

We calculated 3 different measures of alpha diversity, Faith’s Phylogenetic Diversity index [39], Shannon diversity index [40] and Simpson’s index [41], and compared their distribution to determine if there were differences in the richness or evenness of the samples between both groups. No significant differences (p-value > 0.05) between groups were found in any of the indexes tested. In Figure 1-B Shannon diversity index is shown. Hence richness, evenness and phylogenetic diversity were similar between non-UC controls and UC patients. Additionally we observed that the rarefaction curve came to plateau indicating the sequencing depth was sufficient to measure the bacterial community (data not shown). In terms of beta-diversity, we calculated robust compositional Aitchison distances and plotted them with ellipses representing the 80% of confidence interval of each group (Fig. 1-B). Total variability was explained with only three principal components and differences between groups were significant for PERMANOVA analysis (p-value = 0.007 and pseudo-F = 5.34). This indicates that the compositional structure of the community differs between non-UC controls and UC patients.

### Differentially abundant taxa between patients and healthy controls

To study differentially abundant taxa between non-UC controls and UC patients, we used two compositional algorithms: ANCOM, with 0.7 detection rate and ALDEx2. For both algorithm five taxa were significant: *Akkermansia* (*Verrucomicrobia* phylum), with W= 78 and p-value = 0.022, and *Bacteroides* (*Bacteroidetes* phylum), with W= 59 and p-value = 0.017, were more abundant in healthy controls, instead *Collinsella* (*Actinobacteria* phylum), with W= 69 and p-value = 0.008, *Eubacterium* (*Firmicutes* phylum), with W= 67 and p-value = 0.008 and a undefined genus of the family *Coriobacteriaceae*, with W= 55 and p-value = 0.039, more abundant in UC patients (Fig. 1-C). Although the *Anaerotruncus* genus found a trend of overrepresentation in non-UC controls, this value was not statistically significant (W= 50 and p-value = 0.056).

### Functional analysis

To predict the functional capacity of the gut microbiota in non-UC controls and UC patients, we used a full pipeline of PICRUSt2, implemented in QIIME2. PICRUSt uses an extended ancestral-state reconstruction algorithm to estimate which gene families are present and then combines gene families to estimate the composite metagenome. From the data of functional capabilities, we focused primarily on those which were associated with the microbial metabolism. We noticed significant differences in certain metabolic functions in the gut microbiome of non-UC controls and UC patients. In total, 5.561 KEGG orthologs (KO) were predicted for both groups, from which only 157 differ significantly in abundance (p-value < 0.05) (Fig.A). In the figure 2-B was represented the relative abundance of this significant KO annotations grouped by KEGG metabolism. Broadly, gene families associated with Metabolic pathways, Phosphotransferase system (PTS), Galactose metabolism and ABC transporter were overrepresented in UC patients. Instead for non-UC controls, 14 KO annotations were found overrepresented, but these annotations were not part of any specific route.

**Figure 2.**
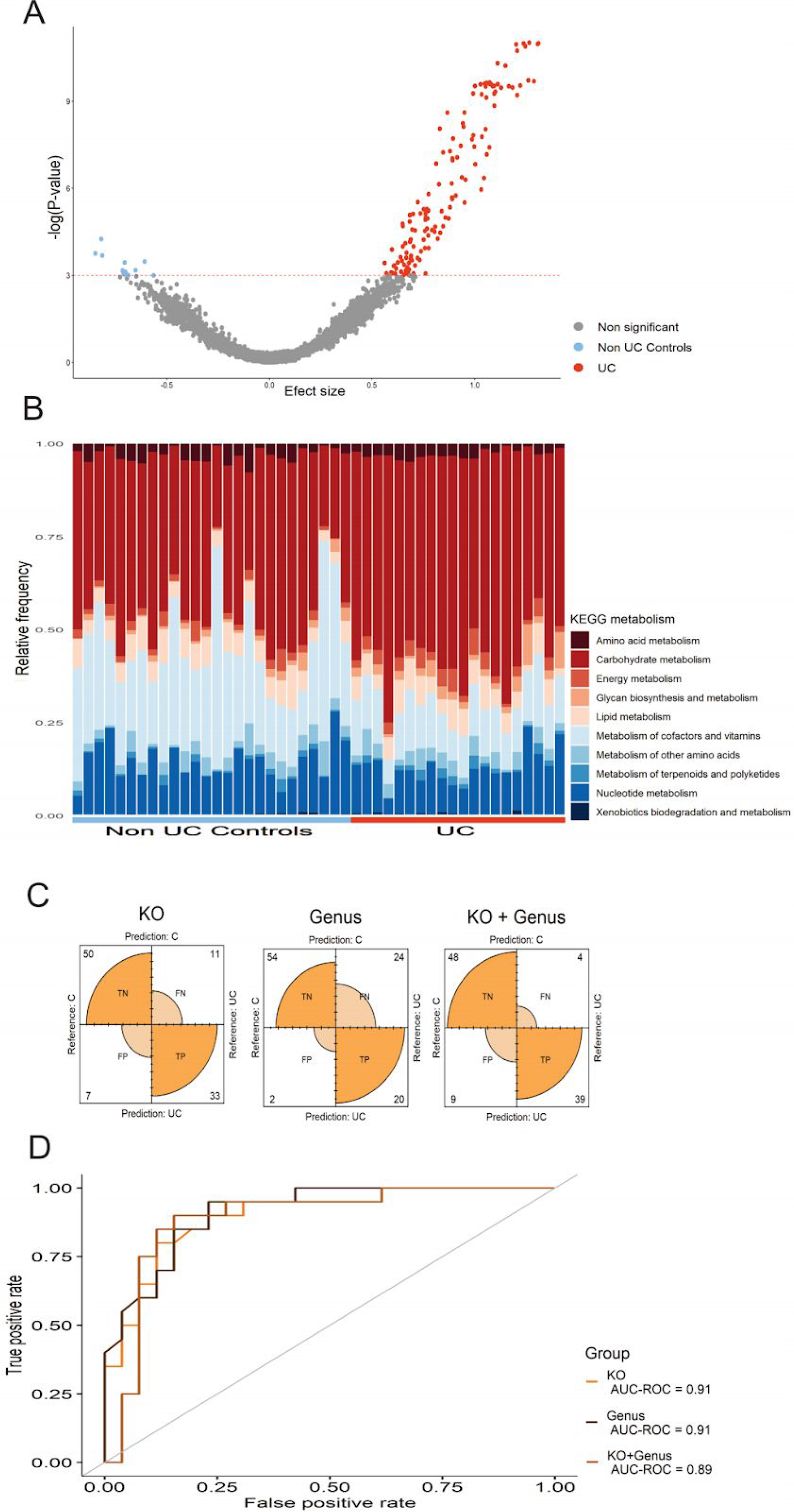
**A)** Volcano plot of the differentially abundant KO between non-UC controls and UC patients. Red dash lines represent significance threshold (p-value = 0.05). Blue represents significant KO more abundant in non-UC controls and red represents significant KO more abundant in UC patients. In gray non significant features are represented. **B)** Significative KO composition grouped by KEGG metabolism in non-UC controls and UC patients. Each column represents one fecal sample and different colors indicate different KEGG metabolism in the microbiota composition. **C)** Confusion matrix about models predictions, y-axis represent true labels and x-axis represent predicted labels. Percentage of the predicted samples are in each corner. **D)** ROC curves and AUC of KO, Genera and KO+Genera models.

### Machine learning performance

Taxonomic and functional annotations were used to classify subjects to their cohort using a machine learning approach. We trained three k-fold cross validation Random Forest classifiers using KO annotations, genus taxonomic level and both (KO + Genera). The overall accuracy was 0.82 for KO model, 0.74 for genus model and 0.89 for KO + Genera model. The correct classification of non-UC controls were 84%, 94% and 87% and correct classification of UC patients was 76%, 46% and 90% respectively (Fig. 2-C). The performance of the models were high with AUCs ranging between 0.89 and 0.91 (Fig. 2-D). The models were also evaluated at the OTU and species level but no better performance was obtained (data not shown).

## DISCUSSION

The present study represents the first report, according to our knowledge, of gut microbiota composition of UC patients in the South America population. As reported previously, IBD has been associated with a decline in the diversity of commensal flora [42–44]. However, with respect to UC patients there is no bibliographical concordance in this point: There is evidence of significant differences in alpha diversity in some populations, as well as there is absence of significant differences between UC patients and their healthy partners who share the same environment [45]. In our study we found no significant differences in alpha diversity between non-UC controls and UC patients. In this sense, it is well known that the microbiome contributes to the host development and physiology stability in frequently encountered environments. However It has been hypothesized that upon encountering a stressful new environment, the microbiome adapts much faster than the host, being their cooperation disrupted, promoting host destabilization and generating reciprocal changes in the host and its microbiome. After chronic exposure to the altered environment, the changes in the microbiome would contribute to the canalization of the altered state. This hypothesis suggests that a new stability of the adapted patterns would be generated, while promoting the variability of the microbiome, which may be beneficial in new stressful conditions. This will allow the host to balance the stability and flexibility of their commensals based on contextual demand [46,47]. This complex dynamic may explain why UC patients did not show significant differences in gut microbiota richness or evenness with non-UC controls (alpha diversity), but microbial compositional structure was more similar within groups than between them (beta diversity). Considering that our patients were under medical treatment, the symptoms of inflammation and bleeding were controlled. In this sense, partial or total remission could define a new microbial state, not identical to the pre-pathogenic one but stable and flexible enough to maintain equilibrium in the new inflammatory context. Additionally, beta diversity changed significantly between groups. This would indicate the key role of microbial structure in this disease [45].

Regarding relative abundance at phylum level, our results found *Firmicutes, Bacteroidetes* and *Proteobacteria* phyla as the most abundant, both in UC patients and non-UC controls. These results are similar to those reported by the literature [48,49]. In IBD, a decrease in *Firmicutes* and an increase in *Proteobacteria* have been reported [50–52]. Strikingly, a trend towards significant abundance of *Firmicutes* was found in UC patients (p = 0.051), being higher than in non-UC controls. There are numerous reports in the literature where this phylum is found overrepresented in inflammatory conditions such as obese subjects [53]. This could be in line with the fact that our patients have average BMI associated with being overweight. However, recent studies have indicated that edge-level associations, such as *Firmicutes/Bacteroidetes* ratio, are not good biomarkers of dysbiosis in the obese patients due to variability among subjects in the same population [54]. Significant relative abundances were also found in *Actinobacteria* and *Verrucomicrobia* phyla (p <0.05). Five genus taxa were differentially abundant using two methodologies: *Collinsella, Akkermansia, Eubacterium, Bacteroides* and an undefined genus of the family *Coriobacteriaceae*. This was congruent with the results found at the genus level, since *Collinsela* belonging to the phylum *Actinobacteria* was significantly higher in UC patients, while *Akkermansia* belonging to the phylum *Verrucomicrobia* was significantly higher in non-UC controls. This last phylum had been previously reported within the general population of Buenos Aires (Argentina) and Santiago de Chile [18,22]. There are several studies where *Akkermansia municiphila* is specifically associated with a healthy gut, as it has been proved its ability to reverse atherosclerotic lesions, to improve the restoration of the intestinal barrier and reduce endotoxin-induced metabolic inflammation [48,55].

As for *Collinsella*, it has been associated with autoimmune diseases such as psoriasis or rheumatoid arthritis, as well as with subclinical inflammation diseases such as obesity and type 2 diabetes [57–60]. Interestingly, IgA-Sec studies (sequencing strategy to study bacteria linked to IgA) showed that *Collinsella aeroficians* is highly coated by this common mucosal antibody in both IBD patients and healthy controls. However, it was only highly coated in IBD individuals for what it was considered to be a colitogenic bacteria [61]. Therefore, *Collinsella aeroficians* could be interpreted as a candidate pathobiont within the ulcerative colitis in our population. Nevertheless, further studies of metagenome shotgun sequencing at the species/strain levels might be useful to evaluate *Collinsella* species distribution associated with our UC patients. In this sense, Ashley R. Wolf et al. show that fructoselysine (FL), a common Maillard reaction product, selectively increases the absolute abundance of *Collinsella intestinalis* but not *Collinsella aerofaciens* in gnotobiotic mice, even though both species can catabolize FL in vitro. *C. intestinalis* grows faster on FL than glucose, while *C. aerofaciens* represses its FL degradation locus in the presence of glucose and grows more rapidly on glucose than FL [62]. The knowledge that *Collinsella intestinalis* metabolizes e-fructoselysine to innocuous products, proposes a key role of gut bacterial species distribution within human health and its role in pathogenic dysbiosis processes. Strikingly, Yilmaz et. al. have recently highlighted the notion that increased *Collinsella*, as well as *Bifidobacterium, Lachnospira, Lachnospiraceae, Roseburia* and *Eggerthella* taxa are related to successful clinical response or remission of anti-TNF therapy in Crohn’s disease [63]. This finding introduces the important concept that the individual’s microbiota might determine the clinical outcome of immunotherapy in IBD. In addition, Fei Sjöberg showed in a pilot study that the duodenal microbiota of children with ulcerative colitis exhibited reduced overall richness, being *Collinsella, Lactobacillus* and *Bacillus* the less frequently detected genera [63,64]. To our knowledge, there are no reports in the literature that *Collinsella sp* is overrepresented in adult UC patients compared to non-UC controls. It would be interesting to highlight if this overrepresentation found in our population is associated with its role as pathobiont or if it is a beneficial microbial state associated with low inflammation grade, as most of our patients were under remission or mild clinical activity.

Other differential genera have also been found in our samples, such as the increase of *Bacteroides* in the non-UC controls, which have previously been associated with a decrease in obesity [65] and irritable bowel syndrome [66]. In addition, the benefits of Bacteroides species such as *B*.*Vulgatus* and *B*.*Dorei* have been reported to reduce the production of intestinal microbial lipopolysaccharides (LPS) [67]. Increased LPS affects the tight junctions of intestinal epithelial cells through Toll Like Receptor 4 (TLR4) [68] in addition to inducing the expression of PC3-secreted microprotein (PSMP), a chemokine expressed in colitis and colonic tumor tissues [69]. Regarding the genus *Anaerotruncus*, not many reports were found at UC patients. Studies in a murine model found this bacterium in the external mucus layer and in the luminal content in control mice [51]. Finally, *Eubacterium* and an unclassified genus of the *Coriobacteriaceae* family were found increased in UC patients. This is consistent with a previously reported association of an unclassified genus of *Coriobacteriaceae* with UC patients [45]. Likewise, *Eubacterium dolichum* is reported to be highly coated with secretory IgA in UC patients [61]. Several studies link the class of *Erysipelotrichi* (where the genus *Eubacterium* is found) with the unhealthy gut and with its colitogenic potential in murine model [61]. In addition, an increase in the abundance of *Erysipelotrichi* was reported in UC patients [70], colon cancer [71] and were correlated with inflammation and immunomodulation [72]. However, more recent studies of intestinal mucosal associated microbiota in UC patients report a decrease in the genus *Eubacterium* compared to healthy controls [73]. Therefore, there is no bibliographic concordance for *Eubacterium* genera and its functional correlation with intestinal disease is far from being fully elucidated.

In addition, we estimated the functional capabilities of the gut microbiota in the studied groups and found some differentially abundant KO annotations within UC patients. Many of these KO annotations are related with carbohydrate metabolism in particular galactose metabolism, a pathway that was described in a strain of Collinsella aerofaciens [74]. This has coherence with genus abundance analysis described formerly. Additionally ABC transporters pathways were also overrepresented in UC patients. In this sense, Davenport et. al. has described that membrane transport (such as ABC) and carbohydrate metabolism were overrepresented in non-inflammatory biopsies of patients with ulcerative colitis [75]. These results reflected in our functional study could be related to Collinsella’s ability to metabolize carbohydrates in-vitro studies, as well as to the clinical low inflammatory remission state that our patients presented in most of the cases [62].

In order to define a robust classifying method in our population, we evaluated the capability of three machine learning models to classify the individuals. These models utilized KO (AUC= 0.91), taxa genus level (AUC= 0.91) and the combination of both (AUC= 0.89). Two of these machine learning models showed similar capabilities, when using KO and genus to distinguish non-UC controls and UC patients. However, the genus model was the worst in identifying UC patients (46% of correct classification). This reinforces the idea of functional compensation in the microbiome community, as models that use KO annotations had better capabilities than taxonomy to distinguish UC patients.

Notably, this is the first report describing the taxonomic composition of intestinal microbiota in Argentinean subjects diagnosed with UC. Although the vast majority of studies of intestinal microbiome composition in different human populations are performed from fecal samples, it is important to note that human fecal microbiota is not a faithful reflection of the cecal or colonic microbiota [14]. Despite this, the use of this sampling methodology is less invasive than endoscopies and biopsies that can lead not only to health but also to ethical problems, since not only sick patients but also healthy volunteers are intervened. Therefore, the study of fecal matter is a limitation that must be considered in the interpretation of the results, but not an exclusion. Additionally, it is important to note that this study is correlational and therefore conclusions related to the pathogenesis of UC disease at these low taxonomic levels are difficult. Even though sample size could be improved, this pilot study contributed to the knowledge of the uncharacterized gut dysbiosis associated with UC patients in the Argentinean population. Similar studies have been conducted in small cohorts [44,76] and the changes observed in gut microbiota of its subjects were clear. Further functional characterization, such as proteomics or metabolomics, as well as longitudinal metagenomic shotgun studies should be performed in South American UC patients. This could improve sampling limitations and consider intestinal mucosa metabolism within our local environmental factors, allowing a better understanding of the role of the gut dysbiosis in these chronic diseases of unknown etiology. In this sense, several publications mention the importance of knowing the microbiome of each region, since it is conditioned by intrinsic local factors [22,77].

In order to analyze the specific microbial signature related to the development of IBD in our region, our results provide a first exploration of the UC-associated microbiota in Buenos Aires (BA) and its metropolitan area, which constitute a megalopolis being the second most populated agglomeration in South America and the southern hemisphere. Possessing this baseline will contribute with the medical specialists to the local treatment of this disease, promoting further studies for the development of new probiotics and prebiotics as well as possible fecal transplants to treat different phenotypes of UC.

## CONCLUSION

This work was focused on the study of the gut microbiota of the metropolitan population of Buenos Aires, Argentina, with the aim of finding biomarkers related to UC. We found no significant differences in the gut microbiota richness or evenness between UC patients and non-UC controls (alpha diversity), but microbial compositional structure was different within groups (beta diversity). *Akkermancia* and *Bacteroidetes* resulted the most representative genera within the controls, while *Collinsella* and *Eubacterium* were found overrepresented in the UC patients. In addition, our results showed that carbohydrates metabolism was preponderant in UC patients, not observing a distinctive metabolism for the healthy non-UC controls. Finally, in order to define a robust classifying method in our population, we evaluated the capability of three machine learning models to classify the individuals. Our results reinforced the idea of functional compensation in the microbiome community, as models that use KO annotations had better capabilities than taxonomy to distinguish UC patients. Overall, this pilot study evidenced gut microbial dysbiosis associated with UC in our population, which contributes as a starting point to future studies of the local microbiome associated with inflammatory pathologies, as well as the possible redefinition of more effective treatments in IBD.

## Data Availability

Data accession

## AUTHOR CONTRIBUTIONS

F.S.B and A.P.S. designed the study. J.C., R.S., C.C., C.M. performed the recruitment of the volunteers. A.R. and S.Q. collected the stool samples and performed the extraction of fecal bacterial DNA. A.P. and A.P.S. processed the raw sequences and performed the bioinformatic and statistical analysis. F.C.L., G.I., A.P.S. and F.S.B analyzed the results. A.R., A.P. and F.S.B. wrote the manuscript. All authors contributed in the manuscript revision.

## CONFLICT OF INTEREST STATEMENT

All authors declare no competing interest.

## ACKNOWLEDGMENTS

The authors thank the “Centro de Investigación, Docencia y Extensión en Tecnologías de la Información y las Comunicaciones” (CIDETIC, http:/cidetic.unlu.edu.ar/), Universidad Nacional de Luján, Luján, Argentina for human and computational resources. We are grateful to Fundación H.A. Barceló, Instituto Universitario de Ciencias de la Salud, CABA, Buenos Aires, for excellent technical support.

## FUNDINGS

This research was supported by the Department of Basic Sciences of the National University of Lujan (UNLu), Florencio Fiorini Foundation, Barcelo Foundation, Argentine Society of Gastroenterology and the National Agency for Science and Technology (PICT2017-2406).

## Notes

### Competing Interest Statement

The authors have declared no competing interest.

### Author Declarations

This study received approval by the Ethics Committees of Hospital Espanol of BA (for non-UC control population) and Hospital Nacional Prof. Alejandro Posadas (for UC patients), according to local regulations and Helsinki declaration. Written informed consent was obtained from all study participants.

